# ESTIMATING UNDERDIAGNOSIS OF COVID-19 WITH NOWCASTING AND MACHINE LEARNING – EXPERIENCE FROM BRAZIL

**DOI:** 10.1101/2020.07.01.20144402

**Authors:** Leandro Pereira Garcia, André Vinícius Gonçalves, Matheus Pacheco Andrade, Lucas Alexandre Pedebôs, Ana Cristina Vidor, Roberto Zaina, Ana Luiza Curi Hallal, Graziela De Luca Canto, Jefferson Traebert, Gustavo Medeiros de Araujo, Fernanda Vargas Amaral

## Abstract

**Background:** Brazil has the second largest COVID-19 number of cases, worldly. Even so, underdiagnosis in the country is massive. Nowcasting techniques have helped to overcome the underdiagnosis. Recent advances in machine learning techniques offer opportunities to refine the nowcasting. This study aimed to analyze the underdiagnosis of COVID-19, through nowcasting with machine learning, in a South of Brazil capital.

**Methods:** The study has an observational ecological design. It used data from 3916 notified cases of COVID-19, from April 14^th^ to June 02^nd^, 2020, in Florianópolis, Santa Catarina, Brazil. We used machine-learning algorithm to classify cases which had no diagnosis yet, producing the nowcast. To analyze the underdiagnosis, we compared the difference between the data without nowcasting and the median of the nowcasted projections for the entire period and for the six days from the date of onset of symptoms to diagnosis at the moment of data extraction.

**Results:** The number of new cases throughout the entire period, without nowcasting, was 389. With nowcasting, it was 694 (UI95 496-897,025). At the six days period, the number without nowcasting was 19 and 104 (95% UI 60-142) with. The underdiagnosis was 37.29% in the entire period and 81.73% at the six days period.

**Conclusions:** The underdiagnosis was more critical in six days from the date of onset of symptoms to diagnosis before the data collection than in the entire period. The use of nowcasting with machine learning techniques can help to estimate the number of new cases of the disease.

## BACKGROUND

The World Health Organization has reported more than 10 million cases of SARS-CoV-2 infection and 500,000 deaths,^1^ a significant part of which had occurred in Brazil. According to the Brazilian Ministry of Health, the country overcame 1,3 million cases and 58 thousand deaths,^2^ what meets the Imperial College London prediction of growing in deaths caused by the COVID19.^3^ Brazil has the biggest number of deaths among the Latino-American countries.^4^ The Lancet has dedicated, recently, an editorial to the political-sanitary disaster that desolate the country.^5^ Despite the already alarming numbers, the editorial^5^ and other studies^6^ had drawn attention to the possibility of a large number of underdiagnosed cases. One of the causes of underdiagnosis is the low testing rate of suspected individuals: 4.71 tests for a thousand habitants.^7^ This rate is much lower than countries like Iceland (184.11), United States (66.76), Chile (30.01), South Africa (16.34).^7^ Dealing with the underdiagnosis is essential so that appropriate actions can be taken to reverse the progression of deaths in the country.^8^

Many countries are using a combination of containment and mitigation activities to stem the progression of SARS-CoV-2 and thus, manage the demand for hospital beds.^9^ Non-pharmacological measures have been shown to be effective in controlling the transmission of COVID-19.^10–14^ They can reduce the impact in the health system, given managers time to organize properly the system. These measures also reduce the need for hospitalization by other conditions that could compete for beds with SARS-CoV-2 patients.^15^ In addition, they increase the chance that a substantial number of people not be infected until a treatment and vaccine be developed.

In outbreak situations, in which rapid changes are common, the actual number of infected cases must be closely monitored. Artefacts variations produced during the monitoring process should be distinguished of the real cases variation.^8^ Among the artefacts are less testing capacity than suspected cases notification, for example. If the number of individuals notified as suspects is much higher than the testing capacity at the present time, this difference can cause an underdiagnosis of the current cases. Data about pathogens transmissibility and exposed population susceptibility, population density and demographic characteristics of the affected population, besides the temporal spatial-distribution of cases and population mobility, can contribute to the correction of such artefacts.^16^

The natural history of the disease, on the other hand, is an important factor in determining the optimal case count update in the frequency monitoring. Rapidly progressing diseases like COVID-19 require daily updates, while monthly updates may be sufficient to others with slower progression, such as HIV / AIDS. A frequent analysis may also be necessary in times when transmissibility is expected to be changing, for example when control actions are initiated, enhanced, or stopped.^16^

Nowcasting approaches try to estimate the number of a given event in the present.^8,10,17^ This strategy has been used to improve surveillance of infectious diseases like AIDS ^18,19^ cholera,^20^ influenza infections^8,21^and, recently, COVID-19.^3,8,17,20,22^ Nowcasting techniques, in general, uses time-series predictions.^23–25^ Recent advances in machine learning techniques offer opportunities to refine the nowcasting of an epidemic behavior.^16^ The main objective of machine learning techniques is to produce a model that can be used to classify, predict, or estimate a phenomenon. This approach is useful in several applications in biomedical research,^26–32^ including concerning COVID-19.^33,34^

Monitoring the impact of non-pharmacological actions is essential to optimize the allocation of scarce resources in non-high-income-countries, like Brazil.^16^ In these, the maintenance of long quarantine periods is even more challenging due to the deficiencies on the social protection system, the economic vulnerability of the population and the large portion of people acting as informal workers. No single set of interventions is appropriate to all contexts owing to the combination of these factors with climatic, demographic and organization issues of each country.^10^ Thus, monitoring on near real time should be a key part of the strategy to couple with SARS-CoV2. Among the challenges for timely monitoring are delays in providing medical care after onset of the symptoms and delays in diagnosis.^8^ It is plausible to assume that these challenges are even greater in non-high-income-countries, with less comprehensive health systems.

To help overcome this challenge, the present study aimed to analyze the underdiagnosis of COVID-19 cases, through nowcasting with machine learning, in a South of Brazil capital city.

## METHODS

### Ethical Considerations

This project was submitted to the Ethics in Research with Human-Beings Council at the Federal University of Santa Catarina to guarantee the alignment with Resolution n° 466/2012 of the National Health Council of Brazil. The research project was approved under CAE n° 33374820.2.0000.0121/2020. We used exclusively secondary and anonymized databases.

### Study Design

The present study has an observational ecological design, using data from notified cases of COVID-19 by the Health Department of Florianópolis, capital of the State of Santa Catarina in southern Brazil from April 14^th^ to June 02^nd^, 2020. Florianópolis has 500,973 inhabitants^35^ and is administratively divided into 49 health regions.^36^ The health regions correspond to the areas covered by the primary health care units. The median time from the date of onset of symptoms of COVID-19 to notification is three days; as well as the time from notification to release of the test result in the city (unpublished data, provided by the Public Health Department of Florianópolis).

We used the random forest^37^ machine learning algorithm to classify the notified cases which had no diagnosis yet, producing the nowcast. To analyze the underdiagnosis, we compared the difference between data without nowcasting and the median of the nowcasted projections for the entire period of analysis and for the period from May 28^th^ to June 2^nd^, 2020. The latter corresponds to the six days from the date of onset of symptoms to diagnosis at the moment of data extraction.

### Definition of Suspected and Confirmed Cases

Notification of suspected cases of COVID-19 within 24 hours is mandatory in Brazil.^38^ From April 14^th^, 2020, Florianópolis adopted the same criterion of notification used by COVID-19 as the criteria used by the Brazilian Ministry of Health: fever accompanied by cough, dyspnea, runny nose or sore throat.^38^ The cases have been confirmed by real time reverse-transcriptase-polymerase-chain-reaction (RT-PCR), serological tests or clinical-epidemiological criteria.

### Data Source and Variables

We used three data sources for the nowcasting, all from the Public Health Department of Florianópolis: 1) anonymized database of suspected and confirmed cases of Florianópolis’ residents; 2) demographic data for the 49 health regions; and 3) traffic data, as a proxy for the movement of people in the municipality.

The following variables were extracted from anonymized database of suspected and confirmed cases: i) diagnostic (confirmed, discarded or missing), ii) sex, iii) age (in years), iv) age groups (under 10 years, from 10 to under 20, from 20 to under 40, from 40 to under 60, from 60 to under 80 and over), v) race (white and not), vi) date of birth, and vii) onset of symptoms. Individual suspected and confirmed cases were extracted on the diagnosis. Data were also extracted on the person’s region of health.

The number of infected people (with a positive diagnosis and less than 14 days of symptom onset) and the rate of infected people per 100,000 inhabitants were calculated for the health regions where each notified person resides. In addition, the following demographic data from these regions were included in the analysis: i) the total number of inhabitants and by sex, ii) the number of persons aged 1 year old, 2 years old and so on up to 100 years old or more, iii) the number of people by race (white, black, yellow, brown, indigenous and ignored), iv) the number of people by years of schooling (from 1 to 17 years completed or more, in addition to literate, non-literate, literate through youth and adult literacy programs and with uninformed schooling), v) total income per household, average income of households, total income of heads of households, average income of heads of households, total income per person and average income per person. The proportion of male people, people aged 60 years old or over, people with non-white race and people with 10 or less schooling time, was calculated as possible indicators of vulnerability.

The average daily traffic in four important avenues in the city was used as a proxy of people’s mobility in the city. We hypothesize that there is a lag between the increase in mobility and the identification of the increase in cases, so we used the average traffic of the day and the average lagged daily until the thirteenth day of the onset of the symptoms of the notified cases.

There was no imputation for missing data.

### Descriptive Analysis

To compare the characteristics of people with a confirmed and discarded diagnosis of COVID-19, t test was used for continuous variables and chi-square for categorical, adopting the p-value < 0.05 as a threshold of statistical significance.

### COVID-19 Incidence Nowcasting with Random Forest

We used the random forest to carry out the nowcasting. The database was initially splitted in the training-validation-test database, formed by cases whose diagnosis (confirmed or discarded) was known; and the prediction database, which had no diagnosis. The training-validation-test database was divided, next, in training-validation database and test database, using 70% and 30% of the data, respectively.

The training-validation basis was subjected to undersampling to improve the sample’s balance as the number of discarded cases was much higher than confirmed. The balanced training-validation database was used to perform the feature selection and hyperparameter tuning. Nested cross validation was performed with 5 folds, both in the inner and outer loop. The feature selection and hyperparametrization were performed simultaneously in the inner loop using a random search to maximize the accuracy. Folds were balanced with respect to the outcome. Table 1 shows the range for feature selection and hyperparameters used.

**Table 1:** Feature selection grid and hyperparameters used to adjust the random forest.

| Action | Hyperparameter | Minimum | Maximum |
| --- | --- | --- | --- |
| Feature Selection | Permutation Importance (percentage) | 0.20 | 1.00 |
| Hyperparameter | Number of trees | 100 | 2000 |
|  | Mtry | 1 | 50 |
|  | Minimum node size | 1 | 10 |
|  | Sample fraction | 0 | 1 |

We analyzed the training and validation results. The model with the best fit was used for classification in the test database. The test database was not submitted to undersampling reflecting the prediction database as close as possible. Finally, the cases were classified as confirmed or discarded, based on the predictions.

We repeated the resampling of the databases, the training and the testing of the algorithms 1000 times to determine the 95% Uncertainty Intervals (UI), the median of accuracy, sensitivity and specificity, in addition to the final classification of cases.

The underdiagnosis was analyzed by the difference between the median of the number of cases predicted by the model (incidence with nowcasting) and the number of the cases diagnosed by the Public Health Department of Florianópolis (incidences without nowcasting). This analysis was carried comparing the entire period and the period from May 28^th^ to June 2^nd^, 2020. The number of cases was also smoothed by a LOESS^39^ regression and the cumulative number, without and with nowcasting, were presented graphically by day of symptom onset.

All analyzes were performed using the software R v.3.6.3. Anonymous scripts and databases are available at: https://github.com/lpgarcia18/underdiagnosis_of_covid_19_cases_in_brazil

## RESULTS

During the analysis period, 3916 individuals residing in Florianópolis were reported as suspects for COVID-19. Among all notified individuals, 603 had a positive diagnosis, 2413 discarded diagnosis and 900 had no diagnosis yet. The association of individual characteristics, health regions and displacement of people with confirmed or discarded cases can be seen at the Table 2 and at the Supplement.

**Table 2:** Association among individual characteristics, health territories and social distance and positive and negative cases of SARS-CoV-2 in Florianópolis, Santa Catarina, Brazil.

|  |  |  |  | p |
| --- | --- | --- | --- | --- |
| FEATURES | POSITIVE (N=603) | NEGATIVE (N=2413) | TOTAL (N=3016) | VALUE |
| INDIVIDUAL NOTIFICATION CHARACTERISTICS |  |  |  |  |
| DATE OF SYMPTOMS ONSET * |  |  |  | < 0.001 |
| Mean (SD) | 2020-04-22 (24.5 days) | 2020-04-29 (17.4 days) | 2020-04-28 (19.2 days) |  |
| DATA OF SUSPITION NOTIFICATION * |  |  |  | < 0.001 |
| Mean (SD) | 2020-05-10 (13.7 days) | 2020-05-06 (13.5 days) | 2020-05-07 (13.6 days) |  |
| GENDER |  |  |  | 0.008 |
| Female | 306 (50.7%) | 1369 (56.7%) | 1675 (55.5%) |  |
| Male | 297 (49.3%) | 1044 (43.3%) | 1341 (44.5%) |  |
| RACE |  |  |  | 0.025 |
| Black | 35 (5.8%) | 127 (5.3%) | 162 (5.4%) |  |
| Brown | 36 (6.0%) | 77 (3.2%) | 113 (3.7%) |  |
| White | 503 (83.4%) | 2079 (86.2%) | 2582 (85.6%) |  |
| Yellow | 29 (4.8%) | 129 (5.3%) | 158 (5.2%) |  |
| Missing | 0 (0.0%) | 1 (0.0%) | 1 (0.0%) |  |
| **AGE** |  |  |  | < 0.001 |
| Mean (SD) | 40.7 (17.2) | 37.6 (18.7) | 38.2 (18.5) |  |
| **AGE GROUP** |  |  |  | < 0.001 |
| Less than 10 | 17 (2.8%) | 214 (8.9%) | 231 (7.7%) |  |
| 10 to 20 | 30 (5.0%) | 116 (4.8%) | 146 (4.8%) |  |
| 20 to 40 | 264 (43.8%) | 1039 (43.1%) | 1303 (43.2%) |  |
| 40 to 60 | 206 (34.2%) | 765 (31.7%) | 971 (32.2%) |  |
| 60 to 80 | 74 (12.3%) | 226 (9.4%) | 300 (9.9%) |  |
| More than 80 | 12 (2.0%) | 53 (2.2%) | 65 (2.2%) |  |
| **HEALTH TERRITORY CHARACTERISTICS** |  |  |  |  |
| **POPULATION** |  |  |  | < 0.001 |
| Mean (SD) | 17357.4 (10731.0) | 15338.4 (8993.6) | 15742.1 (9399.7) |  |
| **MALE PROPORTION** |  |  |  | < 0.001 |
| Mean (SD) | 0.92 (0.06) | 0.93 (0.05) | 0.93 (0.06) |  |
| **INFECTED PEOPLE** |  |  |  | < 0.001 |
| Mean (SD) | 19.0 (8.6) | 16.6 (10.1) | 17.06 (9.9) |  |
| **RATE OF INFECTED PEOPLE** |  |  |  | 0.054 |
| Mean (SD) | 154.5 (137.1) | 141.9 (146.5) | 144.4 (144.7) |  |
| **PEOPLE WHITH MORTE THAN 60** |  |  |  |  |
| **YEARS OLD PERCENTAGE** |  |  |  | < 0.001 |
| Mean (SD) | 0.16 (0.06) | 0.14 (0.05) | 0.14 (0.06) |  |
| **LESS THAN 10 YEARS OF SCHOOLING** |  |  |  |  |
| **TIME PERCENTAGE** |  |  |  | < 0.001 |
| Mean (SD) | 0.6 (0.06) | 0.6 (0.06) | 0.6 (0.06) |  |
| **NON WHITE RACE PERCENTAGE** |  |  |  | 0.045 |
| Mean (SD) | 0.16 (0.10) | 0.17 (0.10) | 0.16 (0.10) |  |
| **MEAN PER CAPITA INCOME– OF THE** |  |  |  |  |
| **HEALTH TERRITORY** |  |  |  | < 0.001 |
| Mean (SD) | R\$ 3685.69 (R\$ 1898.76) | R\$ 3040.96 (R\$ 1609.68) | R\$ 3169.86 (R\$ 1690.92) | |
| SOCIAL DISTANCING |  |  |  |  |
| **TRAFFIC AVERAGE LAG 13 DAYS** |  |  |  | < 0.001 |
| Mean (SD) | 12217.4 (6658.9) | 10462.3 (5032.1) | 10813.2 (5440.9) |  |

The group of individuals with a positive result for SARS-COV-2 had an earlier symptom onset date and later notification dates than individuals with negative results. There was also a difference regarding the distribution according to sex and race between the two groups. The average age among confirmed cases was higher than among discarded cases. There was a heterogeneous distribution of confirmed and discarded cases among the 49 health regions in the municipality. The average number of confirmed cases was higher in regions with a higher average age, proportion of women, average level of education, income, and white people. Most positive cases were observed after seven days of higher average car traffic.

The classification algorithm showed an accuracy of 0.91 (UI 95% 0.83 - 0.97) in the training database, 0.66 (UI 95% 0.62 - 0.69) in the validation and 0.66 (UI 95% 0.62 - 0.69) in the test. The sensitivity and specificity values can be seen in Table 3.

**Table 3:** Values of accuracy, sensitivity and specificity of the classification algorithm.

|  | Train (UI 95%) | Validation (UI 95%) | Test (UI 95%) |
| --- | --- | --- | --- |
| Accuracy | 0.91 (0.83-0.97) | 0.65 (0.62-0.70) | 0.66 (0.62-0.69) |
| Sensibility | 0.91 (0.83-0.98) | 0.65 (0.61-0.69) | 0.65 (0.57-0.79) |
| Specificity | 0.91 (0.82-0.97) | 0.66 (0.61-0.70) | 0.66 (0.60-0.71) |

**Table 4:** Incidence of cases without and with the nowcasting between May and 2 June 2020.

|  | **Incidence – Entire period(UI 95%)** | **Incidence May 28<sup>th</sup> - June 2<sup>nd</sup> (UI 95%)** |
| --- | --- | --- |
| Without nowcasting | 389 | 19 |
| With nowcasting | 694 (496-897) | 104 (60-142) |

The incidence without nowcasting throughout the entire period was 389 new cases. With the nowcasting it was 694 (UI95% 496 - 897). At the period from May 28^th^ and June 02^nd^, 2020, the incidence without nowcasting was 19 new cases and 104 (UI95% 60 - 142) with nowcasting (Table 3). Thus, the underdiagnosis was 37.29% in the entire period and 81.73% in six days from the date of onset of symptoms to diagnosis at the moment of data extraction. The difference in the progression of new cases with and without nowcasting can be seen in Figure 1.

**Figure 1:**
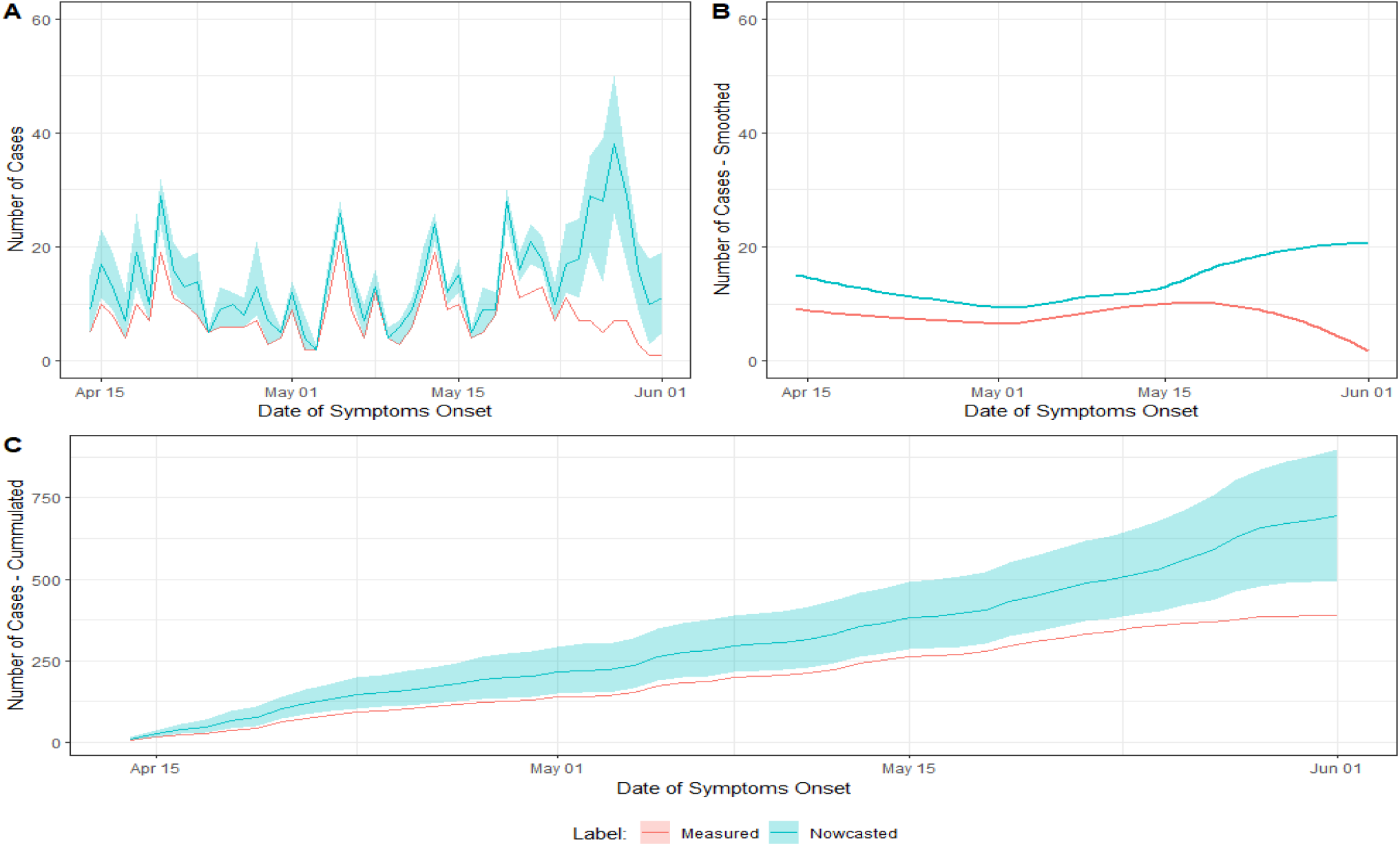
Evolution of the number of new cases with and without nowcasting. A) Number of cases per day of symptom onset; B) LOESS regression of the number of cases per day of symptom onset. C) Accumulated number of cases per day of symptom onset. Legend: the shaded area corresponds to the 95% Uncertainty Interval.

## DISCUSSION

COVID-19 data analysis represents a challenge for statisticians and epidemiologists in non-high-income-countries due to the magnitude of underreporting ^40^ Even so, the number of COVID-19 cases has grown rapidly in Brazil, and the country has the second largest number of cases in the world nowadays.^7^ The city of Florianópolis has, so far, 1686 confirmed cases and 14 deaths caused by SARS-CoV-2.^41^ At the time of this research, the epidemic in Florianópolis reflected the pattern of the introduction of the virus in Latin America, which occurred first in people with higher income, who had traveled to countries where the virus was already present. Thus, a higher average of confirmed cases was observed in health regions with a greater number of white-skinned people, with higher income and schooling patterns. A greater movement of cars in days before the onset of symptoms was also associated with the confirmation of the reported cases. This meets with the evidences regarding the importance of social distance to reduce the transmission of SARS-CoV-2. In a study carried out in Canada, for example, a social distance strategy reduced the number of cases and the transmission of SARS-CoV-2, producing a reduction in intensive care units admissions and deaths^42^.

Maintaining strong measures of social distance for long periods, however, may not be sustainable. These restrictions have already caused a slowdown in the world economy.^43^ Research in non-high-income-countries shows an average 70% fall in income and 30% less in consumption expenses.^42^ Strategies that combine more restrictive periods with moments of relaxation of these restrictions have been identified as ideal for countries with few resources.^44^ Interleaving periods with greater restriction of social contact with periods of relaxation of the restrictions, but with an intensification of testing, case isolation, contact tracking and protection of vulnerable people, can allow a return to the minimum coexistence between people, and the resumption of economic production.^44^

Florianópolis has carried out more than 10,000 tests so far,^45^ that is, more than 20 tests per thousand inhabitants, more than 4 times the national average. Even with this greater number of tests, which should reduce the impact of underdiagnosis in the municipality, it is possible to observe a great disparity between the number of new cases confirmed by the municipality and the one predicted by the nowcasting. The underdiagnosis was more important in the proximal period of analysis. It shows the significance of underdiagnosis in the six days between the date of onset of symptoms and the date of diagnosis prior to the data collection. The underdiagnosis, probably produced by the mismatch between the onset of symptoms and the time of testing, may interfere with the current estimate and future projections of the disease incidence. In this context, the use of machine learning techniques can assist to enable adequate monitoring of the number of new cases and better decision making.^46^

The algorithm performance was better in detecting negative cases (specificity) than positive cases (sensitivity). In this sense, a greater number of false positives are expected compared to false negatives, and the interpretation of nowcasting should take this into account. A greater amount of individual data, such as data related to symptomatology, can improve model sensitivity. Besides, the association of SARS-CoV-2 infection rates with climate issues has been described.^47,48^ The introduction of these data may also assist and should be considered in future studies.

## CONCLUSION

The present study demonstrated the underdiagnosis of cases of COVID-19 in Florianópolis. The underdiagnosis was more important in the period of six days before the data collection than in the entire period, corresponding to an artifact in the monitoring caused, probably, by the greater capacity in notifying than in the testing processes. Adequate new cases estimation on time is essential for monitoring the number of reproductions and decision making in the face of the epidemic. The use of nowcasting with machine learning techniques can help to estimate the number of new cases of the disease.

## Data Availability

Anonymous scripts and all databases used are available at: https://github.com/lpgarcia18/underdiagnosis_of_covid_19_cases_in_brazil

